# Power Outages: An Underappreciated Risk Factor for Children’s Carbon Monoxide Poisoning

**DOI:** 10.1101/2024.07.20.24310120

**Authors:** Alexander J. Northrop, Vivian Do, Nina M. Flores, Lauren Blair Wilner, Perry E. Sheffield, Joan A. Casey

## Abstract

Children’s risk of exposure to carbon monoxide (CO) increases after disasters, likely due to improper generator use during power outages. Here, we evaluate the impact of outages on children’s CO-related emergency department (ED) visits in New York State (NYS). We leveraged power outage data spanning 2017-2020 from the NYS Department of Public Service for 1,865 power operating localities (i.e., communities) and defined all-size and large-scale power outage hours. All-size outage hours affected ≥1% of customers, and large-scale outage hours affected ≥20%. We identified CO poisoning using diagnostic codes among those aged <18 between 2017 and 2020 using the Statewide Planning and Research Cooperative System (SPARCS), an all-payer reporting system in NYS. We linked community power outage exposure to patients using the population-weighted centroid of their block group of residence. We estimated the impact of power outages on CO poisoning using a time-stratified case-crossover study design with conditional logistic regression, controlling for daily relative humidity, mean temperature, and total precipitation. Analyses were stratified by urban and rural communities. From 2017-2020, there were 917 pediatric CO poisoning ED visits in NYS. Most cases (83%) occurred in urban region of the state. We observed an association statewide between all-size and large-scale outages and CO ED visits on the index day and the following two days before a return to baseline on lag day 3. Four hours without power increased the odds of a pediatric CO poisoning ED visit by ≥50% for small-scale and ≥150% for large-scale outages, and associations were stronger in urban versus rural areas. While CO poisoning is a relatively rare cause of pediatric ED visits in NYS, it can be deadly and is also preventable. Expanded analyses of the health impacts of outages and advocacy for reliable energy access are needed to support children’s health in a changing climate

## INTRODUCTION

Carbon monoxide (CO) is a colorless, odorless gas that binds to hemoglobin more avidly than oxygen, which can result in health impacts from mild headaches to death. Major exposure sources include the incomplete combustion of hydrocarbons from vehicle exhaust, gas stoves, and furnaces.^1^ Children may be uniquely vulnerable to CO poisoning due to higher minute ventilation and metabolic demands compared to adults.^2^ Moreover, children’s risk of CO exposure increases after disasters, likely due to improper generator use during power outages.^3,4^ As climate change drives more severe weather events^5^ causing additional outages, we must understand their impact on pediatric CO poisoning. Prior studies evaluated single large-scale outages, which do not represent more frequent, smaller outage exposures nationwide.^6^ Here, we evaluate the impact of outages on children’s CO-related emergency department (ED) visits in New York State (NYS) from 2017-2020.

## METHODS

Power outage data spanning 2017-2020 was obtained through the NYS Department of Public Service for 1,865 power operating localities (i.e., communities), areal units approximately the size of zip codes. We determined the number and percent of customers without power every hour in each community. We defined an hour when ≥1% of customers lacked power as an all-size outage-hour and when ≥20% of customers lacked power as a large-scale outage-hour. The daily sum of outage hours in a community represented the exposure. We stratified communities as urban and rural for subgroup analyses due to differential exposure patterns and characteristics between these groups.^7^

We leveraged block group level outcome data from the Statewide Planning and Research Cooperative System (SPARCS), an all-payer reporting system in NYS. Included cases were CO poisoning among those aged <18 between 2017 and 2020 using a T58 *International Classification of Diseases-10* (*ICD*-10) primary diagnosis (Toxic effects of carbon monoxide).^8^ We linked community power outage exposure to patients using the population-weighted centroid of their block group of residence. An individual is considered exposed if they experienced ≥ 1 outage-hour on either the day of their healthcare encounter or during the two days prior.

We used a time-stratified case-crossover study design with a conditional logistic regression, matching on the day of week, month, and year to control for variables not expected to significantly change during the study period. The models controlled flexibly for daily relative humidity, mean temperature, and total precipitation. These rasterized weather data were obtained from the North American Land Data Assimilation System Phase 2^9^ and aggregated to the community level. We calculated the population attributable fraction (PAF), the proportion of children’s CO ED visits due to all-size and large-scale outages.^10^ All analyses were conducted on R (version 3.6.3; version 4.3.2).

An institutional review board approved the study (#19-00355).

## RESULTS

From 2017-2020, there were 917 pediatric CO poisoning ED room visits in NYS. Most cases (83%) occurred in urban regions (**Table 1**). Most exposed and unexposed cases were non-Hispanic (NH) white children. Among those with a CO ED visit, there was a higher percentage (16%) of Hispanic children who were exposed to outages than NH Black (9%) or (14%) NH white children.

**Table 1:** Demographics Information of Carbon Monoxide Emergency Department Visits in New York State Among Children (<18 years old) Exposed and Unexposed to Power Outages. “Exposed” refers to children residing in a community with ≥1% of customers without power on lag day 0-2 of their presentation to the emergency department for carbon monoxide poisoning. Race and ethnicity data was restricted to three categories due to low counts of other identities, and thus, do not sum to the total number of cases. Race and ethnicity categories represent a social construct without a biological basis. Abbreviations – NH: Non-Hispanic.

|  | **Exposed Cases** | **Unexposed Cases** | **Total Cases** |
| --- | --- | --- | --- |
| **No. (%)** | No. (%) | No. (%) | No. |
| **Total Cases** | 124 (14) | 793 (86) | 917 |
| **Region** |  |  |  |
| Rural | 14 (9) | 145 (91) | 159 |
| Urban | 110 (15) | 648 (85) | 758 |
| **Age** |  |  |  |
| Age <1 | 11 (12) | 80 (88) | 91 |
| Age 1-5 | 37 (13) | 256 (87) | 293 |
| Age 6-10 | 37 (17) | 176 (83) | 213 |
| Age 11-17 | 39 (12) | 281 (88) | 320 |
| **Sex** |  |  |  |
| Female | 59 (13) | 390 (87) | 449 |
| Male | 65 (14) | 403 (86) | 468 |
| **Race/Ethnicity** |  |  |  |
| NH Black | 19 (9) | 193 (91) | 212 |
| Hispanic | 29 (16) | 147 (84) | 176 |
| NH White | 58 (14) | 348 (86) | 406 |

We observed an association statewide between all-size and large-scale outages and CO ED visits (**Figure 1**). Four hours without power increased the odds of a pediatric CO poisoning ED visit by ≥50% for all-size and ≥150% for large-scale outages. Associations were stronger in urban versus rural areas. The population attributable fraction of these outages was 4.8 and 2.6 percent of statewide CO ED visits using the all-size and large-scale thresholds, respectively.

**Figure 1:**
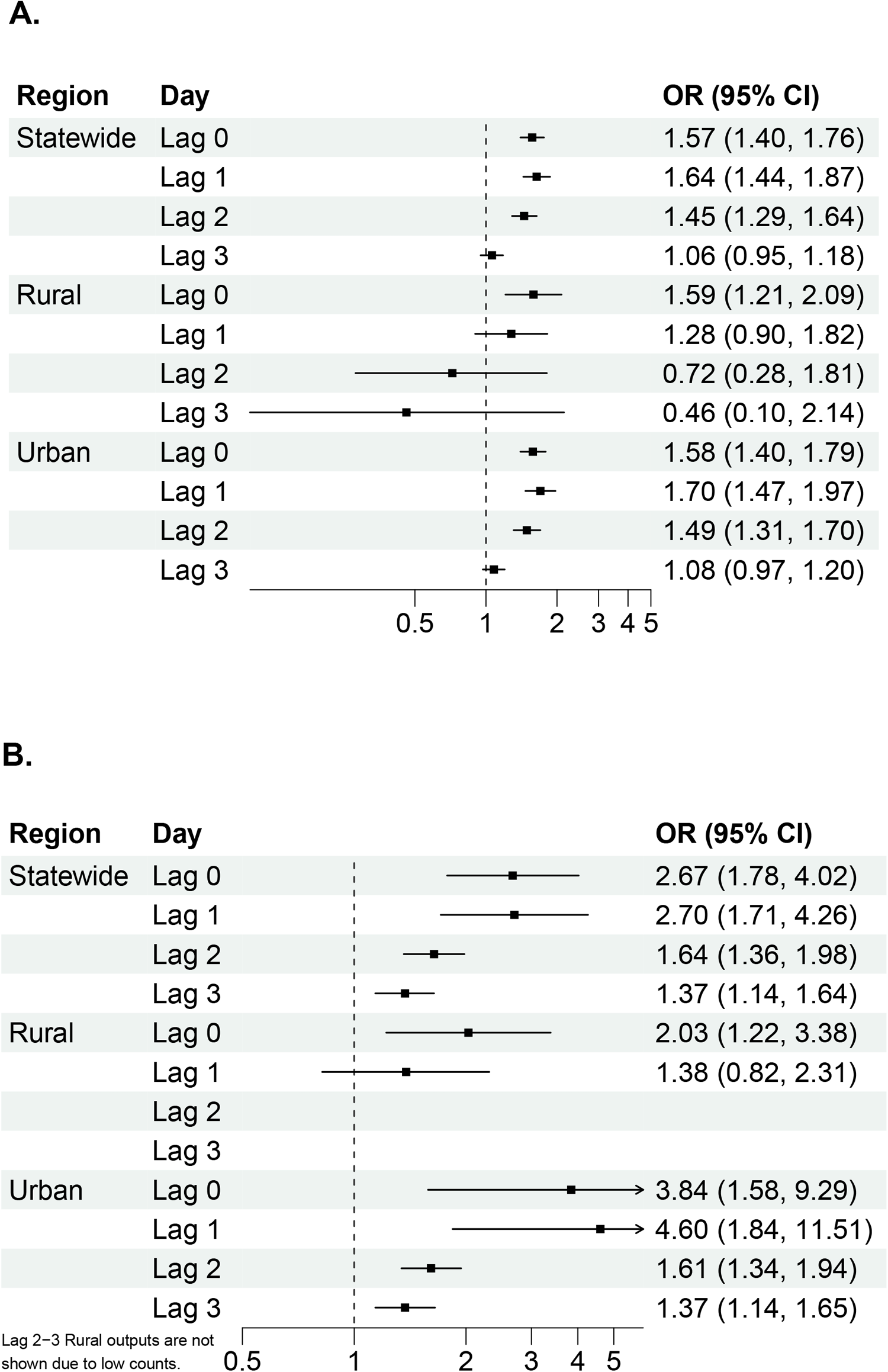
Odds Ratios: Outages and Carbon Monoxide Emergency Department Visits by Thresholds and Urbanicity. Adjusted odds ratios represent the odds of a carbon monoxide emergency department visit for a four-hour increase in community outages at the **A)** all-size outage threshold (≥1% of the community without power) and **B)** large-scale outage threshold (≥20% of the community without power).

## DISCUSSION

While CO poisoning is a rare cause of pediatric ED visits in NYS, it is both frequently deadly and preventable. Our findings leverage exposure data with high spatiotemporal resolution to demonstrate that even smaller-scale outages, captured by our all-size metric, contribute to pediatric CO poisoning ED visits, building upon prior work identifying large-scale outages as a risk factor for CO poisoning.^3^

Outage data was an aggregate community exposure, so we cannot determine if an individual case was truly exposed, potentially biasing our results to the null. Due to low power, we could not evaluate response disparities by social determinants of health. Expanded analyses of the health impacts of outages and advocacy for reliable energy access is needed to support children’s health in a changing climate.

## DISCLAIMER

This publication was produced from raw data provided by the New York State Department of Health (NYSDOH). However, the conclusions derived and views expressed herein are those of the authors and do not reflect the conclusions or views of NYSDOH. NYSDOH, its employees, officers, and agents make no representation, warranty, or guarantee as to the accuracy, completeness, currency, or suitability of the information provided here.

## Data Availability

Power outage data and related code are available upon request. The meteorological data used in the analyses are publicly available. Health data from the analyses may be available upon SPARCS approval.

https://ldas.gsfc.nasa.gov/nldas

https://www.health.ny.gov/statistics/sparcs/access/

## Abbreviations

CO: Carbon monoxide
ED: Emergency department
ICD-10: *International Classification of Diseases-10*
NH: Non-Hispanic
NYS: New York State

